# The Unreliable Judges: Assessing Reproducibility and Self-Preference Bias of LLMs as Free-Text Evaluators

**DOI:** 10.64898/2026.06.15.26355670

**Authors:** J I Alvarez-Arenas, D Jimenez-Carretero, D Mañanes, F Sanchez-Cabo

## Abstract

Large Language Models (LLMs) are transforming clinical practice and research, but their adoption requires rigorous evaluation. While human assessment is ideal, its cost has driven the widespread use of LLMs as evaluators. We introduce an open-source reciprocal framework comparing 71 human experts against six LLMs. AI evaluators show a strong self-preference bias, yet neither group reliably identified whether a response was human- or AI-generated. AI scores correlated with surface features such as length and lexical diversity, whereas human scores did not. By probing the evaluator’s hidden states and applying targeted steering, we show that verbosity is a major causal driver of the bias. Moreover, shuffling question–response pairings shows that long responses keep high scores even when they no longer answer the question, whereas short ones do not, demonstrating that AI judges reward verbosity largely independently of content alignment. Finally, API-based and batch inference inflate stochasticity, underscoring the need for controlled deployment.

## Introduction

Artificial intelligence (AI) is becoming increasingly important in healthcare and biomedical research ^1^. Recent advances, especially in large language models (LLMs), have made it possible to answer clinical questions and assist in clinical decision-making ^2,3^. However, due to the prohibitive computational resources required for training, foundational LLMs often suffer from outdated knowledge and, despite recent advancements, remain prone to hallucination. Evaluation of LLMs is hence a key endeavour, particularly for its implementation in the clinical area. These evaluations must be extensive but also comprehensive and of high quality. Human expert reviews offer contextual judgments and are the preferred source of evaluation, but they are expensive both in terms of money and time 4 and are sometimes inconsistent ^4^. On the other hand, automated metrics like BLEU ^5^ make large-scale evaluation possible, though they often miss the meaning of the complex generated content ^6,7^. The idea of using language models as evaluators—often referred to as “LLM-as-a-Judge”—has recently emerged as a compromise solution offering both efficiency and a reasonable degree of contextual understanding ^8–13^. Another important issue is that most benchmarks currently available are based on multiple-choice questions ^14,15^. While this framework can serve to assess the rigor of the answers, they don’t truly assess the ability of the model to create a coherent and concise text to give an answer, nor the verbosity of the answers. All in all, we and others ^13^ have found a lack of methodological frameworks to systematically evaluate the quality, rigor and conciseness of text-free answers generated by LLMs, what compromises its application on highly regulated fields such as health.

In this work, we propose a systematic framework to assess the accuracy, completeness, and clarity of answers generated by LLMs. We generated over 490 cardiology-related questions and their corresponding answers using a cardiology-specific Retrieval-Augmented Generation (RAG) system built upon a corpus of 650,000 cardiovascular research abstracts from PubMed ^16^, human experts also answered these questions. These answers were evaluated by 71 human experts but also by six smaller-scale language models, including standard and Chain-of-Thought (CoT) models. Moreover, we evaluated a dimension often overlooked in clinical AI validation: the degree of stochasticity of model outputs. While setting the temperature of the model to 0 is thought to ensure consistent predictions, we demonstrate that inference architecture, specifically API-based versus local execution, and sequential versus batch processing, introduces variability that persists even with fixed random seeds. This has critical implications for clinical deployment, where consistency is a fundamental safety requirement.

Our results confirm through quantitative and objective metrics that the use of “LLM-as-a-Judge” poses serious concerns. Moreover, they also provide new insights about the mechanisms underlying Generative AI.

## Results

### Open-source reciprocal evaluation framework

Figure 1a schematically depicts the reciprocal evaluation framework used in our benchmark. In brief, a set of 500 unique clinical questions with their corresponding answers were generated, covering various aspects of clinical practice and research in cardiology. These questions were generated using a cardiology-specific Retrieval-Augmented Generation built upon Llama 3.1 8B ^17^ and including 650,000 PubMed cardiology-specific abstracts ^16^. Each generated question underwent a quality filtering step based on groundedness, relevance to biomedical research, and context independence, what yielded a curated benchmark of 269 high-quality questions.

**Figure 1:**
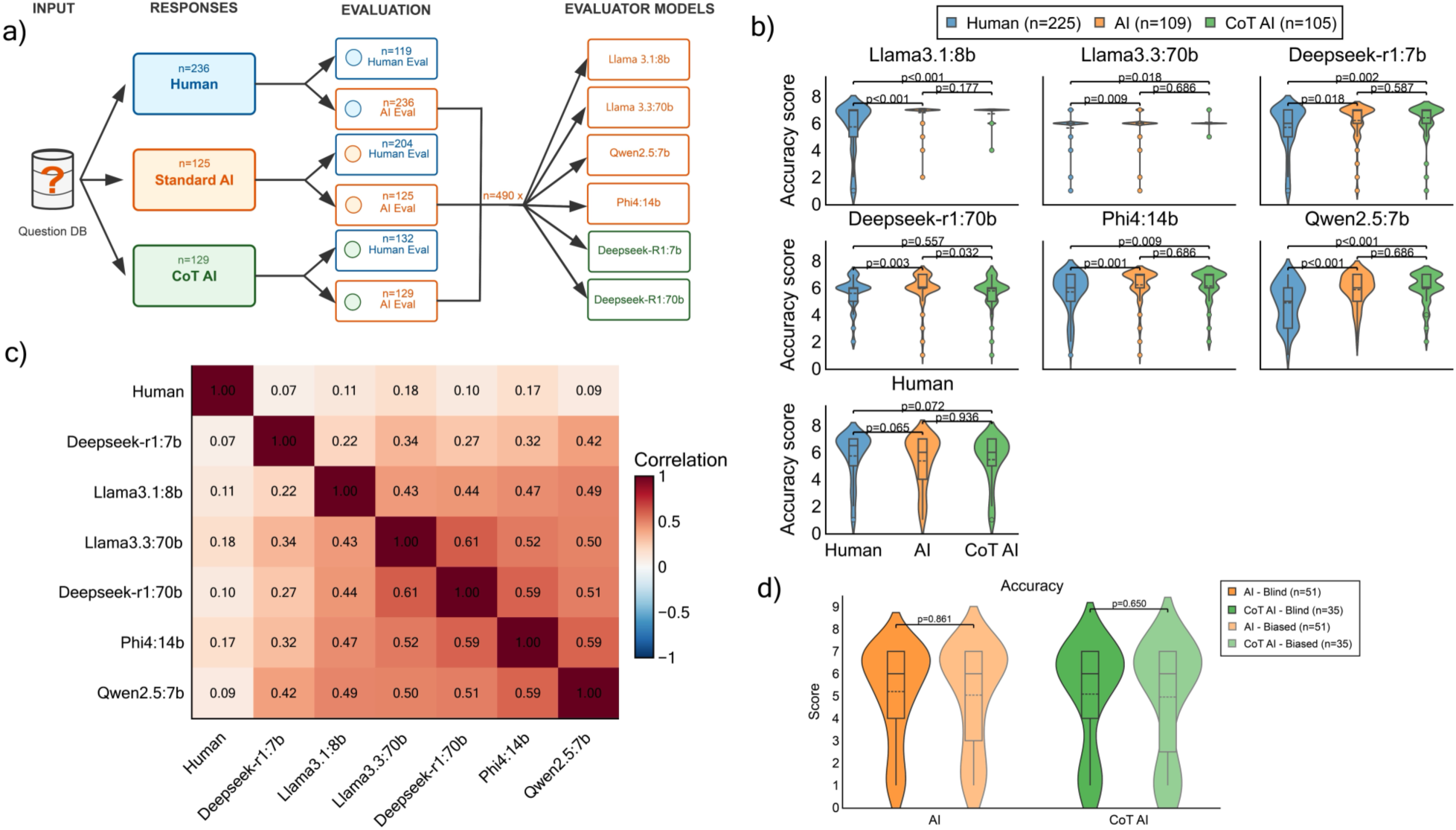
Unbiased evaluation of AI and human generated answers. (a) Scheme of the experimental design. Clinical questions drawn from a curated cardiovascular question database were answered by three sources: human experts (n=236), a standard AI model (Qwen2:7b, n=125), and a Chain-of-Thought AI model (DeepSeek-R1:7b, n=129). Each response was independently evaluated by both human evaluators and by six LLM evaluators (Llama3.1:8b, Llama3.3:70b, Qwen2.5:7b, Phi4:14b, DeepSeek-R1:7b, DeepSeek-R1:70b), yielding a total of 490 evaluated responses per model. “I don’t know” responses were discarded from subsequent analyses. (b) Distribution of accuracy scores across evaluators. Each evaluator (6 LLMs and Humans) is represented in a panel. The violin plots show the distribution of the accuracy scores ranging from 1 to 7, assigned by each evaluator to each type of answer: Human responses (n=225, blue), AI responses (n=109, orange) and CoT AI responses (n=105, green). Violin plots over boxplots show the median, IQR, outlier and frequencies for each distribution. Benjamini-Hochberg adjusted Mann-Whitney p-values are shown. (c) Hierarchical clustering analysis of pairwise correlations between evaluator accuracy assessments. Heatmap displaying Spearman correlation coefficients between accuracy scores assigned by different evaluator types across the same set of responses. Evaluators include six AI-based models (Llama3.1:8b, LLama3.3:70b, Deepseek-r1:7b, Deepseek-r1:70b, Phi4, Qwen2.5) and human expert assessments. (d) Comparison of accuracy scores assigned by humans under source-informed (biased) or blinded responses for AI-generated (orange) and CoT AI-generated (green) conditions. No significant differences were observed between blind and biased evaluations for either response type (p=0.861 and p=0.650, respectively), indicating that human evaluators maintained consistent scoring regardless of disclosed response origin.

71 biomedical researchers and cardiologists were recruited and categorized according to their primary area of expertise. For each question answered by experts, two additional responses were generated by the same RAG system ^16^ but now using two small models different from the one used to generate the questions: one standard LLM (Qwen2-7B ^18^) and one Chain-of-thought model (DeepSeek-R1-7B ^19^). In total, one human answer and two AI-based answers were available to evaluate different aspects of free-text AI-generated responses. From the original 71 researchers recruited to answer the questions, 53 took part in the human evaluation process, completing 455 assessments overall. Human evaluators were randomly assigned batches of 6–9 responses per session, balanced to include approximately one third from human experts, one third from standard AI models, and one third from CoT AI models, with source attribution always concealed. Participants were free to complete as many evaluation sessions as they wished or to stop at any point, which naturally resulted in an uneven distribution of evaluations across responses. No response was evaluated more than once by the same participant. In total, we had 204 evaluations of standard AI-generated responses, 132 of CoT answers, and 119 evaluations of human answers.

All 490 answers generated by humans, AI and CoT were also evaluated independently by 6 LLMs: LLaMa3.1:8b, Llama3.3:70b, Deepseek-r1:7b ^19^, phi4:14b ^20^, deepseek-r1:70b ^19^ and Qwen 2.5:7b ^18^. This yielded a total of 2,940 AI-generated evaluations. Further details can be found in the Methods section. The questions, answers and evaluations are freely available at: https://huggingface.co/datasets/Jialvareza/cardio_evaluations AI evaluators are biased towards AI-generated answers

To assess evaluation objectivity, human evaluators and LLM evaluators scored each answer with a number ranging from 1-7 for three response attributes: accuracy, completeness, and clarity. All evaluators were blinded to the origin of the responses.

Unbiased evaluation results show that all LLMs used as evaluators preferred answers generated by standard AI over human generated answers, suggesting a pattern of optimistic self-assessment and reinforcement bias in their evaluations (Figure 1b). The same was true for the answers generated by CoT models, with the only exception of DeepSeek 70B which scored human answers similarly to CoT answers. Human evaluators scored human responses only slightly better than AI answers (p=0.065). Similar results were obtained for completeness and clarity scores (Supplementary Figure 1).

To further explore the discrepancy in the judgement of humans and AI-based evaluators, we computed the pairwise Spearman’s correlations across the accuracy scores generated by the five AI models and the human evaluators (Figure 1c). Human evaluations show weak correlation with most AI evaluators (r=0.07–0.18), suggesting distinct evaluation criteria between human experts and automated systems. The evaluations of the different AI models were only moderately correlated (r=0.22–0.59).

After this unbiased analysis, we told evaluators the origin of the next 3 responses they had to evaluate. Interestingly, there is no significant change in the evaluations after knowing if the answer had been generated a standard AI or a CoT AI (Figure 1d).

### Neither humans nor AI-based evaluators identify the origin of the answer

To quantify the ability of humans and AI evaluators to differentiate human from AI-generated responses, classification metrics were calculated for each evaluator (Figure 2a). Human evaluators achieved an F1-score of 0.558 (accuracy=0.512, precision=0.843, recall=0.417), indicating a near-random overall classification performance despite high precision. This result suggests that when humans labelled a response as human-generated, they were often correct, but they failed to identify most AI responses as such (recall=0.417), systematically classifying them as Human-generated. On the other hand, AI evaluators performed substantially worse across all metrics. F1-scores ranged from 0.115 (Deepseek-r1:7b) to 0.324 (Llama3.1:8b), with recall values as low as 0.063 for Deepseek-r1:7b, indicating that AI models almost entirely failed to recognise AI-generated responses. These results reinforce the hypothesis that AI-generated responses have reached a level of perceived quality that makes them largely indistinguishable from human responses, at least in the context of this evaluation. Moreover, they reveal that AI evaluators, despite their strong preference for AI-generated content in scoring, paradoxically lack the ability to reliably identify it.

**Figure 2:**
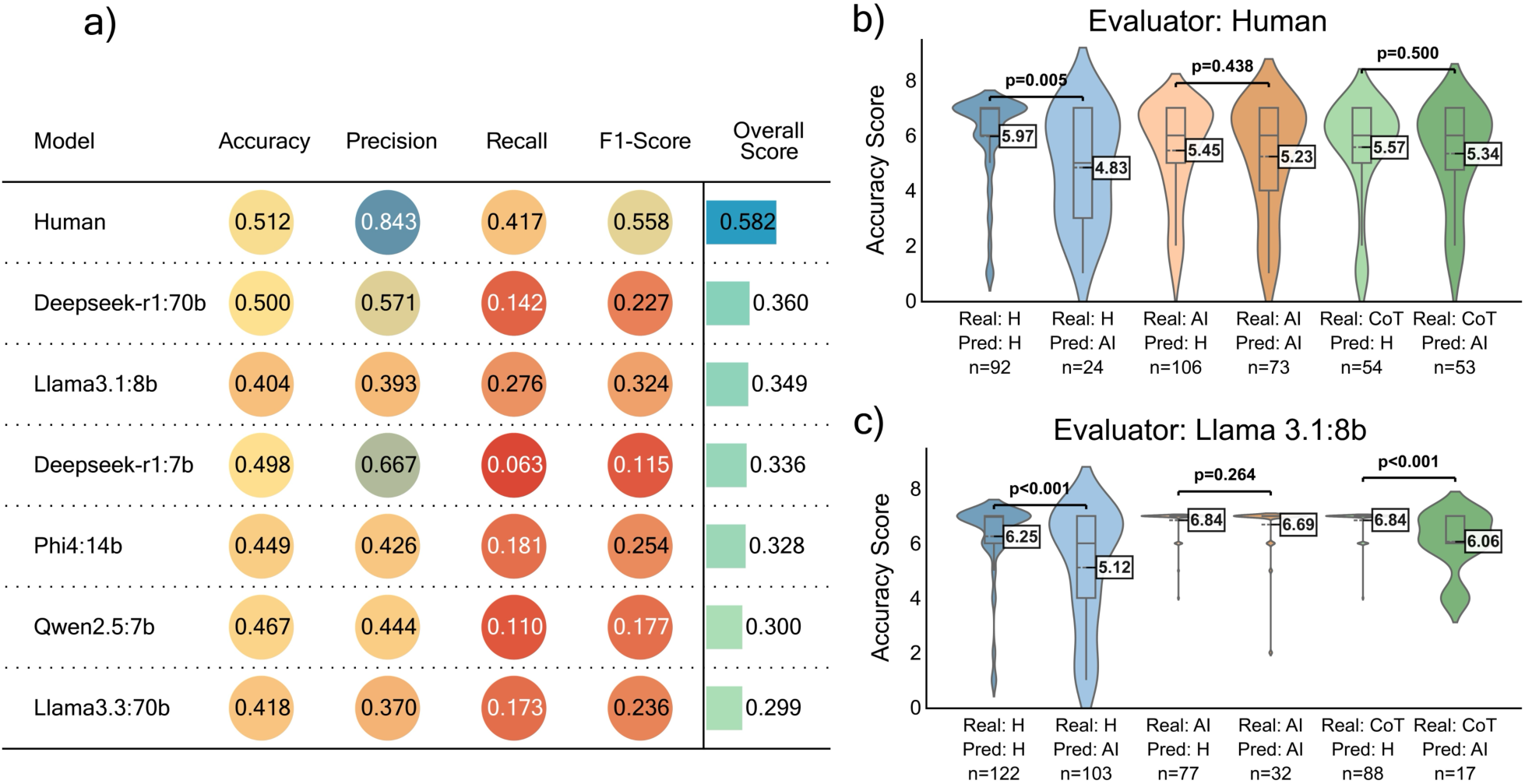
Classification performance of human and AI evaluators in identifying the origin of responses. (a) Table showing accuracy, precision, recall, F1-score, and overall score for human evaluators and six AI models. The overall score is computed as the unweighted mean of the four classification metrics. Evaluators are ranked by overall score. (b-c) Distribution of accuracy scores given by different types of evaluators (human (b), standard AI (c)) distinguishing between actual origin and that predicted by the evaluator. It is observed that both human and AI evaluators tend to give higher scores to responses they perceive as coming from humans. However, the AI shows a clear bias toward its own responses, regardless of perceived attribution.

To conciliate these results with the self-preference exhibited by AI evaluators, we stratified the accuracy scores given by human (Figure 2b) and by standard-AI evaluators (Figure 2c) based on the guessed origin of the answers. Human evaluators scored significantly higher human answers perceived as generated by humans (p=0.005). In contrast, humans scored very similarly all AI-generated answers, independently of their perceived origin. Similarly, AI evaluators scored significantly higher (p<0.001) human texts perceived as humans (Figure 2c). Interestingly, the standard AI evaluator also scored significantly higher those answers generated by CoT model and perceived as human generated, with no significant differences based on origin perception for standard-AI generated answers, although not all models follow this pattern (Supplementary Figure 3). As observed in Figure 1b, the accuracy score for the standard AI evaluator was significantly higher for AI generated answers than for human answers (p<0.001).

### AI evaluators reward length rather than content

To dissect the source of the self-preference bias, we first compared linguistic features between response types and computed Spearman correlations between each feature and the accuracy scores assigned by every evaluator (Supplementary Figure 4). Human responses were significantly shorter, while standard AI and CoT AI responses were longer and more noun-heavy (Supplementary Figure 2). Standard AI evaluators (Llama3.1:8b, Qwen2.5) showed strong statistically significant positive correlations with length-related features — word count (r=0.26, 0.35), sentence count (r=0.32, 0.41), lexical diversity (r=0.28, 0.36), and named-entity count (r=0.33, 0.41) — whereas human evaluators did not correlate with any of them (r ∈ [−0.03, 0.08]). To test whether length was merely a confounder or an actual driver of the bias, we stratified scores by response-length tertiles (Figure 3a). Small AI model evaluators (less than 14b parameters) systematically rewarded longer responses, whereas 70B models and humans did not.

**Figure 3:**
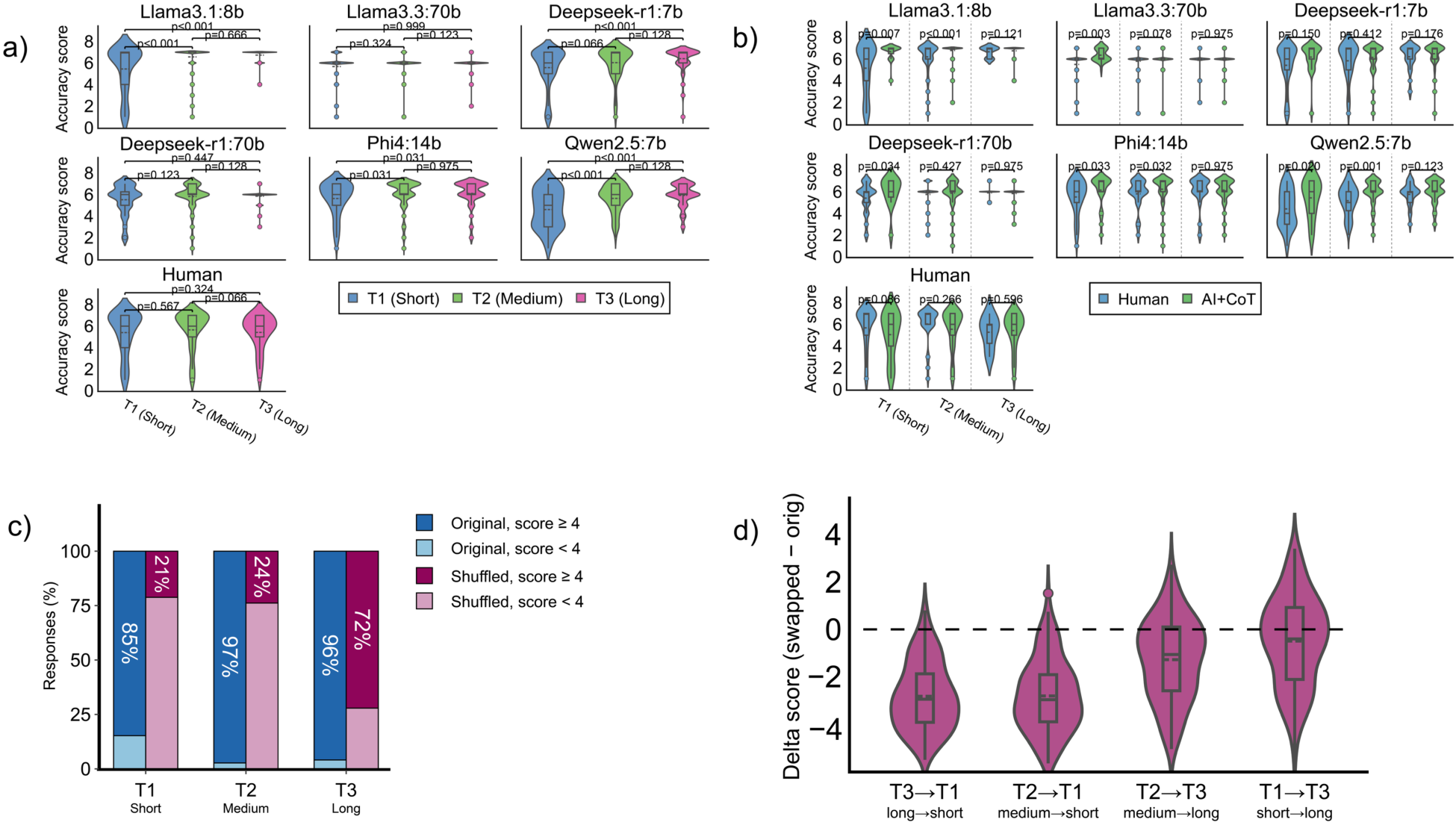
Length, not content, drives the AI-evaluator self-preference bias. (a) Accuracy score distributions for each evaluator stratified by response-length tertile (T1 short: <=83 tokens, T2 medium: 84-159 tokens, T3 long: >=160 tokens); small AI evaluators (<14B) reward longer responses, whereas 70B models and humans do not. (b) Comparison of the accuracy score given by different models to Human (blue) vs pooled AI+CoT AI (green) responses, stratified by the length of the answer in tertiles. (c) Proportion of responses receiving a good accuracy score (≥4) on the original question–response pairs and after shuffling responses within each length tertile, holding response length within the same tertile. Good rates are given to 85% (T1 answers), 97% (T2) and 96% (T3) of the original question/answer pairs, falling after within-tertile shuffling to 21% and 24% for short and medium responses but only to 72% for long ones. Llama 3.1:8B, all 445 Human and AI responses. (d) Change in accuracy score between the original question/answer pair and a shuffled one, across different tertiles of length. Δ = score(shuffled) − score(original). Ti -> Tj denotes the original question in Ti been replaced by one random answer from Tj. (T3→T1, mean Δ=-2.82; T2→T1, mean Δ=-2.81; T2→T3, mean Δ=-1.28: T1→T3, mean Δ=-0.49), Violin plots over boxplots show the distribution, median and IQR; the dashed line marks Δ=0; p-values from Mann–Whitney U tests with Benjamini-Hochberg correction.

Stratifying further by response origin within each length tertile (Figure 3b) revealed that the AI-vs-human gap is concentrated in short responses (T1) and narrows in long ones (T3) — not because the evaluator becomes fairer, but because nearly all long responses are scored high regardless of origin, suggesting that length confounds authorship. To test directly whether the evaluator’s score reflects the semantic match between question and response, we shuffled the question–response pairings under two complementary schemes.

We first shuffled within each length tertile — re-scoring every response against a different question of comparable length — and recorded the proportion of responses still receiving a good score (≥4) (Figure 3c). Shuffling significantly reduced the rate in all three tertiles but the magnitude of the effect decreased sharply with response length. For short responses (T1) the fraction of responses with a good accuracy score collapsed from 85% on the original pairs to 21% after shuffling, and for medium responses (T2) from 97% to 24%. For long responses (T3), however, 72% of the shuffled answers still got a good accuracy score (>=4), even though the answer was unrelated to the question.

To test directly whether it is the length of the response what drives this leniency, we then shuffled across tertiles, replacing each response with one of a different length, and measured the per-response change in score Δ = score(shuffled) − score(original) (Figure 3d). As expected, breaking the question–response link lowered the score in all scenarios — a sanity check confirming that the evaluators are not scoring at random and do retain some sensitivity to content. However, the difference ranged between 0.49 and 2.82, a small drop considering that shuffled responses are totally unrelated to their assigned questions. Importantly, the magnitude of the change in accuracy significantly correlated with changes towards shorter answers (Δ T2→T1=-2.81; Δ T3→T1=-2.82), while changing a true short answer by an unrelated long answer almost didn’t change the accuracy score (Δ T1→T3 =-0.49). In fact, almost 50% of the questions for which the answers were interchanged by random but longer ones (41.5% for T1→T3 and 25.9% for T2→T3) were scored better than the actual answers. In other words, the penalty for answering the wrong question is largely waived — and occasionally reversed — if the substitute is longer: length substantially offsets the cost of irrelevance, and this content-independent leniency grows with length.

### Biases are linearly encoded and causally steerable

Having established behaviourally that length governs the evaluator’s score, we next asked how this bias is represented inside the model and whether it can be causally manipulated. Using Llama3.1:8b — the small open model with the strongest bias — as a testbed for representation analysis, we extracted residual-stream hidden states at every layer for the evaluated responses, teacher-forcing the judge prompt with its response up to the token immediately preceding the accuracy score. These hidden states were used to train logistic-regression probes with 5-fold cross-validation for robustness for three labels of interest: authorship (Human vs. AI/CoT), evaluator-assigned accuracy (scores 6 or more vs 4 or less), and response verbosity (top vs bottom length quartile) ^21,22^.

All three concepts were linearly decodable with near-perfect cross-validated area under the receiver operating characteristic curve (AUROC) (Figure 4a). Authorship was separable at layer 6 (AUROC=1.00), accuracy emerged later, peaking at layer 18 (AUROC=0.95) and verbosity was already separable at layer 1 (AUROC=1.00). At their respective peak layers, the three probe directions were almost orthogonal (cosine similarity < 0.15 throughout, Figure 4b), indicating that authorship is not a relabeled length signal but rather a separate stylistic axis.

**Figure 4:**
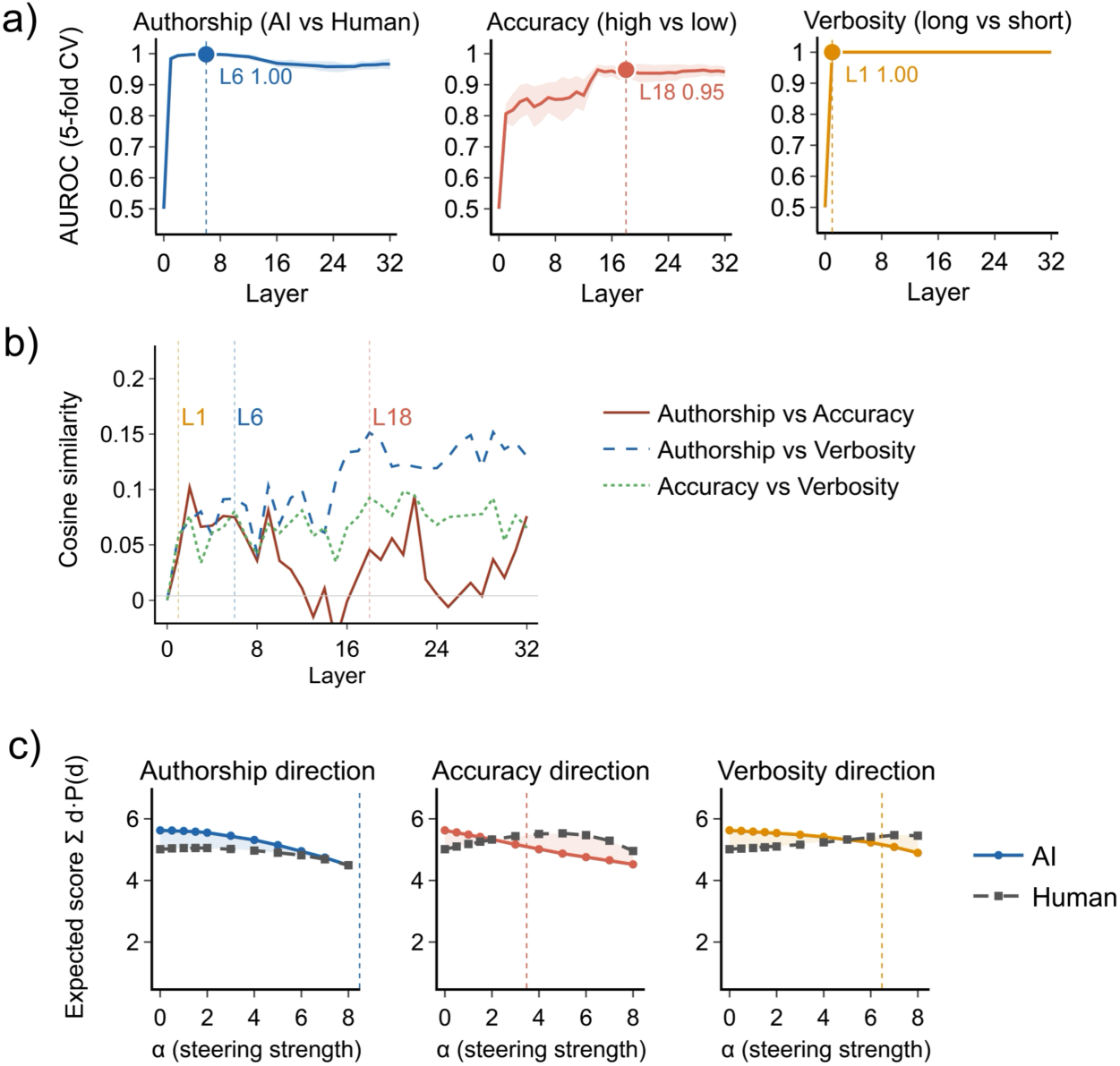
Mechanistic interpretability of the length bias in the Llama3.1:8b evaluator (n=445 responses; AI=109, CoT AI=109, Human=227). (a) AUROC of layer-wise logistic-regression probes (5-fold cross-validation) trained on the residual-stream activation at the token immediately preceding the Accuracy digit, for three labels: authorship (AI vs Human), evaluator accuracy (top vs bottom tertile of Llama3.1:8b scores) and verbosity (top vs bottom tertile of response length). Shaded bands and dashed lines indicate fold variability and peak layer, respectively. Verbosity is separable at layer 1 (AUROC=1.00), authorship at layer 6 (AUROC=1.00) and accuracy peaks at layer 18 (AUROC=0.95). (b) Pairwise cosine similarity between probe directions across layers; vertical lines mark the peak layers from (a). All pairs remain below 0.15, indicating that authorship, accuracy and verbosity are encoded along approximately orthogonal directions. (c) Asymmetric activation steering at the fifteenth layer: for AI prompts the residual stream is projected against the probe direction, for Human prompts along it, with strength α. Curves show the expected accuracy score E[d] = Σ d·P(d) computed from the next-token logits at the Accuracy position. Steering along the verbosity direction is sufficient to close the AI-vs-human accuracy gap, whereas steering along the authorship direction lowers the AI score without symmetrically raising the Human score.

To establish causality, we then applied asymmetric activation steering at the fifteenth layer (see methods) of the model — projecting AI activations against the probe’s direction and Human activations along it — and measured the change in the model’s expected accuracy score read directly from the logits (Figure 4c). Steering along the verbosity direction was sufficient to eliminate the AI-vs-human accuracy gap, an effect of comparable magnitude to steering along the accuracy direction itself. In contrast, steering along the authorship direction lowered the AI score but did not raise the Human score, indicating that authorship and length-related style explain partially overlapping but non-identical fractions of the bias.

### Intra-Model variability and reproducibility in LLMs are controlled only under sequential local inference

Reproducibility is key for the implementation of LLMs in many fields, particularly in the clinical area and in biomedical research. To characterize the stochasticity of LLM-based evaluators under realistic deployment conditions, we generated 5 independent replicas of the complete evaluation set for each of the six models across six inference configurations varying execution environment, request scheduling, and seed control.

Our reproducibility analysis revealed systematic differences in inference consistency across computational conditions (Figure 5). Models with less than 70B were deployed in BF16 precision, with API models and 70B models quantized to 4-bit using AWQ to enable single-GPU inference—a design choice driven by the recognition that tensor parallelism across multiple GPUs introduces additional sources of variability through non-deterministic collective operations and varied floating-point operation ordering.

**Figure 5:**
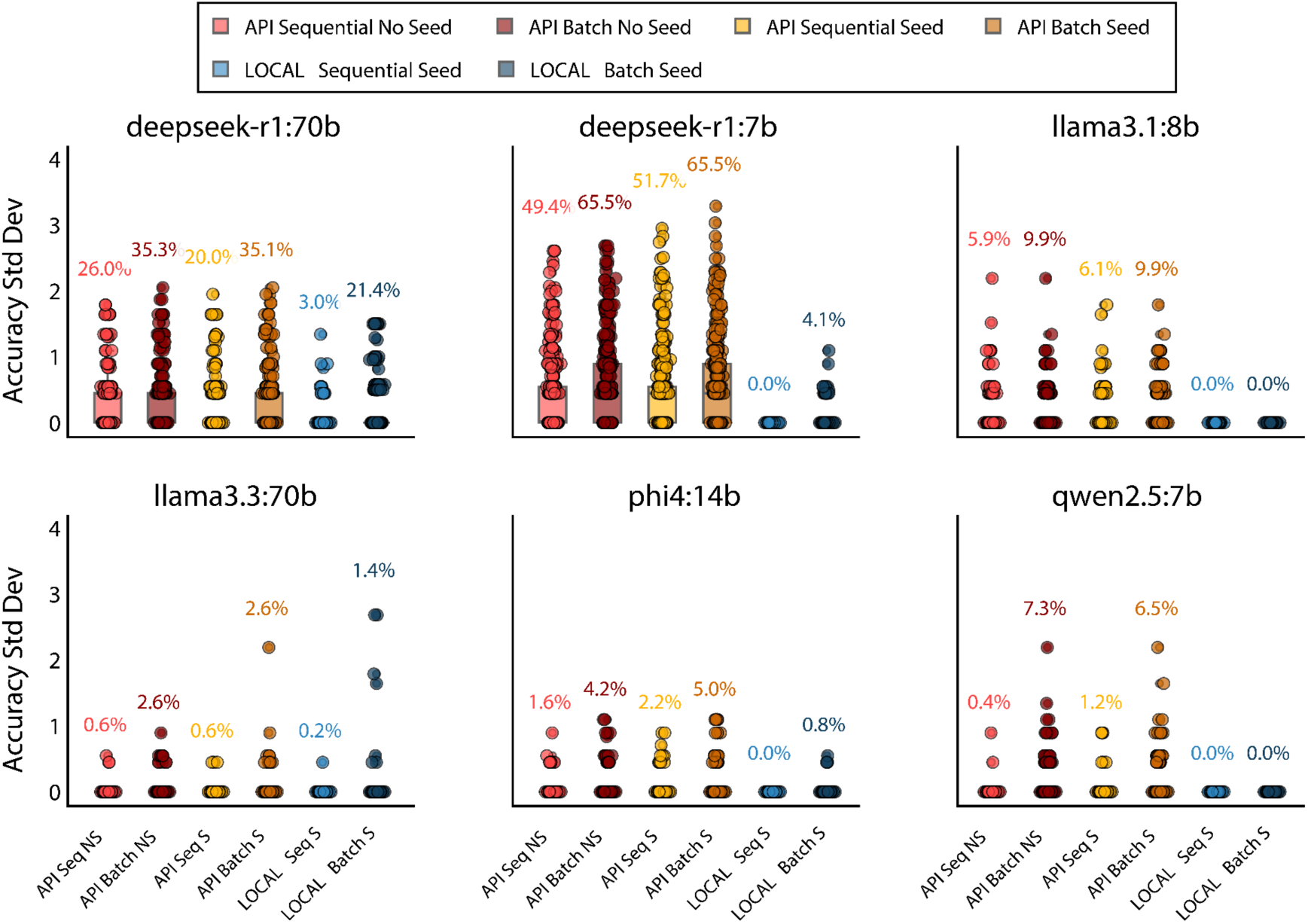
Reproducibility analysis across computational conditions and model architectures. Box plots show the distribution of accuracy standard deviations calculated across 5 independent replicas for six evaluator models under different inference conditions. Each condition is defined by execution environment (API vs. LOCAL), processing mode (Sequential vs. Batch), and seed specification (with/without fixed seed). All evaluations used temperature=0. Numbers above each distribution indicate the number of std different than zero expressed as a percentage, quantifying relative variability across replicas. API models are quantized to 4-bit to run in one single GPU. Local models are BF16 except 70B that are quantized to 4-bit to run in 1 GPU.

Standard models (llama3.1, llama3.3, phi4, qwen2.5) exhibited remarkably low variability across all conditions, with LOCAL sequential execution achieving near-perfect reproducibility (0-0.2% predictions with std dev ≠ 0). In contrast, CoT models demonstrated substantially higher variability: DeepSeek-R1:7b showed 49-66% variable predictions under API conditions, while DeepSeek-R1:70b exhibited 20-55%. This variability persisted even under local sequential execution with fixed seeds (0-4.1% and 3-21% variable predictions, respectively), indicating that the extended reasoning chains introduce compounded sampling uncertainties that interact with numerical precision limitations inherent to BF16 format (7-bit mantissa, non-associative floating-point arithmetic).

API-based inference consistently produced higher variability than local execution across all models, with batch processing amplifying this effect (up to 65.5% variable predictions for deepseek-r1:7b). Notably, seed specification provided minimal benefit in API contexts, suggesting that non-deterministic GPU operations and concurrent request handling introduce variance beyond application-level control.

Critically, even local sequential execution with exclusive GPU access and fixed seeds cannot guarantee absolute zero variability due to fundamental floating-point arithmetic limitations in BF16 precision and 4-bit quantization—where the non-associative property of addition operations (a+b)+c ≠ a+(b+c) under finite precision creates irreducible rounding errors that accumulate differently across execution contexts ^23^. This represents a fundamental trade-off between memory efficiency (enabling consumer GPU deployment) and perfect reproducibility.

## Discussion

Our investigation reveals critical challenges in clinical LLM evaluation and deployment that extend beyond traditional performance metrics. Most current benchmarks evaluate multiple-choice accuracy rather than open-ended text generation, and those assessing free text rely exclusively on AI evaluation, approaches that harbour systematic biases with serious implications, as our findings demonstrate.

Human experts proved unable to reliably distinguish AI-generated responses from human content, achieving near-random classification accuracy. While this demonstrates maturity in text generation capabilities, it fundamentally complicates validation workflows that depend on human judgment. More problematic is the systematic self-preference bias observed in AI evaluators, who assigned significantly higher scores to AI-generated content regardless of its quality.

Behaviourally, the shuffling experiments (Figure 3) show that small open-weight AI judges reward length itself rather than content: when question–response pairings are broken, long responses retain a passing score in 72% of cases even though they no longer answer the question, whereas short responses collapse from 85% to 21%; and on cross-tertile swaps the change in score is governed by the length of the substitute response, not by whether it matches the question. Mechanistically, probing and steering analyses on the same evaluator (Llama3.1:8b) trace this leniency to a causal substrate: response length is linearly encoded from the very first layer, authorship from layer 6, and the evaluator-assigned accuracy only crystallizes around layer 18 (Figure 4), and steering activations along the length direction was sufficient to close the AI-vs-human accuracy gap, whereas steering along the authorship direction did not symmetrically restore human scores. Because AI-generated answers are systematically longer than human ones, this length-driven leniency directly feeds the self-preference bias. This covert mechanism operates below the model’s capacity to articulate it and is largely invisible to human-centred validation workflows.

Our reproducibility analysis reveals a critical but often overlooked dimension in AI validation. Chain-of-Thought models exhibited substantially higher prediction variability, with 20-66% of predictions showing non-zero standard deviation under API-based inference, compared to 0.6-10% in standard architectures. This variability persisted even under supposedly deterministic conditions (temperature=0, fixed seed), as demonstrated by deepseek-r1 models showing 20-52% variable predictions in API sequential seeded conditions versus near-zero variability (0-3%) for local sequential execution. API-based inference showed variable predictions in 0.4-51.7% of cases even with sequential requests and fixed seeds, while batch processing amplified this effect further (reaching 65.5% for deepseek-r1:7b). This arises from non-deterministic GPU operations (CUDA kernel scheduling, parallel execution ordering, floating-point non-associativity exacerbated by reduced precision formats) and concurrent request handling that introduce variance beyond application-level control. Only local sequential execution with exclusive GPU access and seed control achieved near-deterministic reproducibility (0-0.2% variable predictions across standard models), a deployment constraint impractical for most clinical applications and still subject to numerical precision effects when using quantized models. The combination of architectural factors (CoT reasoning chains), deployment constraints (API-based inference), and numerical precision trade-offs (model quantization for GPU memory constraints) creates a reproducibility crisis invisible to current evaluation frameworks. This issue becomes particularly relevant in high-stakes domains such as health, where even small inconsistencies can have practical implications for patient safety.

CoT models offer enhanced reasoning capabilities that may better align with decision-making processes, yet our evaluation study found that they did not show improvement over standard models in expert perception, despite the theoretical advantages of explicit reasoning chains. This suggests that the reproducibility cost is not offset by meaningful performance gains in specialized medical domains where standard models are already well-calibrated.

The QA database generated in this study, comprising multi-model responses, dual reciprocal evaluations from AI and experts, and systematic replica predictions, provides an empirical foundation for developing more reliable validation frameworks. This resource enables direct comparison of evaluation methodologies and quantification of the tradeoffs between different architectural choices in clinical contexts.

Future work should explore methods to mitigate self-preference bias in AI evaluation, develop metrics better aligned with expert judgment, and validate these findings across different domains and tasks. Higher-precision inference strategies such as FP32 may offer a path toward balancing memory efficiency with reproducibility. Ultimately, we hope this work encourages the community to treat the robustness of LLM-based evaluation as a first-class concern—favouring deterministic readouts from the logits where sampling can be avoided, reporting the hardware and inference configurations that shape these results, and assessing reproducibility across deployment conditions—so that as language models become embedded in scientific evaluation, the conclusions we draw from them rest on solid ground.

## Methods

### Question and answer generation

A set of 500 unique clinical questions covering various aspects of clinical practice and research in cardiology were generated through a two-phase pipeline. First, a cardiology-specific Retrieval-Augmented Generator built upon Llama 3.1 8B and including 650,000 PubMed cardiology-specific abstracts 17 was prompted to produce concise, search-engine-style factoid questions, together with an answer and an APA-formatted bibliographic reference. Second, each generated question underwent a quality filtering step based on three independent criteria — groundedness, relevance to biomedical research, and context independence — all scored on a 1–5 scale by the same LLM (Llama 3.1 8B), retaining only questions that achieved a minimum score of 4 across all three dimensions and a manual filtering after that. This process yielded a curated benchmark of 269 high-quality questions. 71 biomedical researchers and cardiologists were recruited and categorized according to their primary area of expertise: cardiac electrophysiology, cardiac imaging, cardiovascular genetics, general cardiology, genomics and transcriptomics, immunology, proteomics, and other related specialties. Altogether, these 71 experts answered at least one out of the 269 high-quality questions, generating a total of 236 answers for 129 questions. For each question answered by experts, two additional responses were generated by our RAG system 17 now using two small models different from the one used to generate the questions: one standard LLM (Qwen2-7B) and one Chain-of-thought model (DeepSeek-R1-7B). In total, one human answer and two AI-based answers were available to evaluate different aspects of free-text AI-generated responses. Both human annotators and AI models were explicitly instructed to respond with “I don’t know” when unable to provide a confident answer to a given question. These responses were subsequently excluded from all downstream analyses. In total, 11 “I don’t know” responses were filtered from the human set, 16 from the standard AI, and 24 from the CoT model.

### Unbiased Evaluation

From the original 71 researchers recruited (respondents), 53 took part in the human evaluation process, completing 455 assessments overall. Human evaluators were randomly assigned batches of 6–9 responses per session, balanced to include approximately one third from human experts, one third from standard AI models, and one third from CoT AI models, with source attribution always concealed. Participants were free to complete as many evaluation sessions as they wished or to stop at any point, which naturally resulted in an uneven distribution of evaluations across responses. No response was evaluated more than once by the same participant. In total, we had 204 evaluations of standard AI-generated responses, 132 of CoT answers, and 119 evaluations of human answers, a total of 455 human evaluations. All 490 answers generated by humans, AI and CoT were also evaluated independently by 6 LLMs: LLaMa3.1:8b, LLaMa3.3:70b, Deepseek-r1:7b, phi4:14b, deepseek-r1:70b and Qwen 2.5:7b. This yielded a total of 2940 AI-generated evaluations.

### Biased Evaluation

Once the unbiased evaluation process was completed, humans and AI were informed about the source of the answer for 111 responses, which corresponded to 63 different questions. A total of 149 biased evaluations were recorded.

### Evaluation metrics

To evaluate the quality of the responses, each human or AI evaluator should grade each response with a number between 1 and 7 ^24^ with the following criteria, used also in similar studies such as GROUSE and G-EVAL ^25,26^:

- Accuracy: Assesses the factual correctness of the information provided and whether it is grounded in evidence.
- Completeness: Measures whether the response includes all relevant information available in the reference documents.
- Clarity: Evaluates the appropriateness and readability of the language used, as well as the ability to directly address the question without introducing irrelevant content.

While all three metrics were collected from both human and AI evaluators, the present study focuses primarily on Accuracy, as it is the least subjective of the three criteria and provides the most informative summary of system performance in a clinical context. Results for Completeness and Clarity are reported in the Supplementary Material (Figure 1a, 1b).

### Linguistic analysis

To characterise structural and stylistic differences between response types, linguistic features were extracted from all 490 responses using the spaCy NLP library ^27^ and the textstat package. Features were grouped into three categories:

- Count features: word count, sentence count.
- Surface features: average word length, average sentence length, unique word ratio, and length-normalised type-token ratio (TTR) as a measure of lexical diversity ^1^.
- Structural features: part-of-speech distributions (verb, noun, adjective, adverb, and pronoun ratios), named entity count, connector phrase count, mean syntactic dependency distance as an index of sentence processing complexity ^2^, Flesch Reading Ease score, and Automated Readability Index (ARI).

Differences between evaluators across response types (human, standard AI, CoT AI) were assessed using pairwise Mann-Whitney U tests with Benjamini-Hochberg correction. Spearman correlation coefficients between each linguistic feature and the accuracy scores assigned by human and AI evaluators were computed using bootstrap resampling (1000 iterations with 50 samples per iteration) to estimate confidence intervals (95% CI). Correlations were considered statistically significant when the confidence interval excluded zero. This approach allowed us to identify the textual properties driving each evaluator type’s judgements while accounting for uncertainty in the correlation estimates. Length tertiles (T1, T2, T3) used in Figure 3a–b were defined on the token-level response length, splitting the dataset at the 33rd and 66th percentiles within the pooled response set.

### Question–response shuffling

To test whether the evaluator’s score depends on the semantic match between question and response, we shuffled the question–response pairings under two complementary schemes. In the within-tertile scheme, each response was re-paired with a different question whose original response fell in the same length tertile (cut-offs at the 33rd/66th percentiles of the pooled response set: T1 ≤83 tokens, T2 84–159 tokens, T3 ≥160 tokens; tertile means 45, 124 and 234 tokens, respectively), holding response length approximately constant; we compared the proportion of responses scoring ≥4 between matched and shuffled pairings within each tertile. In the across-tertile scheme, each response was replaced by one drawn from a different length tertile, and we measured the per-response change Δ = score(swapped) − score(original) for each swap direction. In both schemes the judge prompt was rebuilt with the mismatched pair and the accuracy score recomputed from the logits.

### Hidden-state extraction

Mechanistic interpretability analyses were carried out on Llama3.1:8b loaded with TransformerLens at BF16 precision on a single H100 GPU. For each of the 445 evaluated responses (after filtering “I don’t know” answers with a different regex method than above) we constructed the full judge prompt (system instructions, question and response) followed by the model’s own JSON header up to the token that immediately precedes the Accuracy digit, and ran a forward pass without sampling. The residual-stream activation at the score-token position (i.e. the last token before the accuracy digit) was stored for all 33 layers (embedding + 32 transformer blocks), giving a tensor of shape (445, 33, 4096) per sample position.

### Linear probes

At every layer we trained a binary logistic-regression probe on the residual-stream hidden states (scikit-learn LogisticRegression; L2 penalty, C=1, lbfgs solver, balanced class weights, features standardised in-fold with StandardScaler). The probe is parameterised by a weight vector w ∈ ℝ^4096 — one coefficient per dimension of the layer’s hidden state — plus a scalar intercept, so it defines a single hyperplane in activation space whose normal we interpret as the direction along which the target concept is linearly encoded. Performance is reported as the mean AUROC over 5 stratified folds (StratifiedKFold, seed=42).

Three binary labels were used:

- Authorship: 1 = response generated by AI or CoT-AI, 0 = Human response.
- Accuracy: 1 = response scored at or above 6 by Llama3.1:8b, 0 = scored at or below 5.
- Verbosity: 1 = response token length above the 75th percentile, 0 = below the 25th percentile (token length measured as the offset between the response-start and score-token positions in the judge prompt).

For geometric analysis the weight vector learned in the standardised space was first rescaled back to the original activation space by dividing element-wise by the per-feature standard deviation, and then unit-normalised. This rescaled, unit-normalised vector is what we refer to as the probe direction. Pairwise cosine similarity between probe directions was computed at every layer for all three label pairs (Figure 4b).

### Causal localisation of the bias by activation patching

To justify the layer at which steering interventions were applied, we performed activation patching on Llama3.1:8b to localise where in the network the self-preference bias is causally encoded. For each of 200 matched question pairs we built a clean prompt (the same question paired with the AI response, which typically yields a high score) and a corrupted prompt (same question paired with the Human response, which yields a lower score). Both prompts were tokenised and forwarded through the model. For every layer L in turn, we copied the clean residual-stream activation at the score-token position into the corresponding position of the corrupted forward pass and measured the resulting change in the score metric Δ(L) = M(corrupted with layer L patched from clean) − M(corrupted), where M is the difference in log-probability between high-score tokens ({“6”, “7”}) and low-score tokens ({“1”, “2”, “3”, “4”, “5”}) at the Accuracy position. A large positive Δ(L) indicates that layer L alone carries enough information to flip the model’s score from “low” back to “high”, i.e. that the bias-relevant computation has already taken place by that layer. Because TransformerLens additionally exposes the attention-output and MLP-output streams, we ran the same patching sweep on those two components, yielding three patching curves (residual stream, attention out, MLP out) for the 32 layers of the model. The patching effect on the residual stream was negligible up to layer 14, rose sharply from layer 15 onwards, and approached saturation by the final layers. Patching the attention and MLP outputs individually produced markedly smaller effects, indicating that the bias is not localised in a single component but accumulates in the residual stream across the second half of the network. On the basis of this analysis, we used layer 15 — the first layer with a clearly non-zero patching effect — as the default steering layer.

### Activation steering

Following the representation-engineering framework of Zou et al. ^21^ we used the per-layer probe directions as steering vectors. At inference time we registered a forward hook on the residual stream at the fifteenth layer and applied the projection h ← h − αv for AI-labelled prompts and h ← h + αv for Human-labelled prompts, with α ∈ [0, 8]. The hook was active for every token in the sequence. After steering we read the next-token log-probabilities at the Accuracy position and computed the expected score Σ d·P(d) over the digit tokens d ∈ {1,…, 7}. The asymmetric formulation (subtract on AI, add on Human) was chosen so that residual-norm effects of the projection are absorbed symmetrically by both groups.

### Reproducibility analysis under deterministic conditions

To assess the stochasticity of LLMs, we conducted a systematic variability study across all six evaluator models (llama3.1:8b, llama3.3:70b, deepseek-r1:7b, deepseek-r1:70b, phi4:14b, qwen2.5:7b). For each model, we generated 5 independent replicas of the complete evaluation set under six inference configurations varying execution mode (API vs. local), request scheduling (sequential vs. batch), and seed control (fixed vs. unspecified):

- API Sequential, No Seed: Inference requests sent sequentially to Ollama API without seed parameter
- API Sequential, Seed: Sequential API requests with fixed seed parameter
- API Batch, No Seed: Concurrent API requests without seed parameter
- API Batch, Seed: Concurrent API requests with fixed seed parameter
- LOCAL Sequential, Seed: Local execution with sequential processing and fixed seed
- OCAL Batch, Seed: Local execution with batch processing and fixed seed

All conditions used temperature=0. For each condition and model, we calculated the standard deviation of accuracy scores across the 5 replicas to quantify inference variability. This design allowed us to isolate the effects of execution environment, processing strategy, and seed specification on output consistency.

### Q&A Database and code availability

This study has enabled the construction of a structured cardiovascular question-answering database comprising human expert responses, multi-model AI generations, and dual evaluation tracks (clinical expert and AI assessments). This resource represents a valuable benchmark for the scientific community, providing a foundation for evaluating future specialized AI models for text-free answer in for biomedical research. Unlike existing benchmarks that rely on multiple-choice formats, our database captures the complexity of open-ended clinical discourse with expert-validated quality assessments across accuracy, completeness, and clarity dimensions. To facilitate reproducibility and enable further research, all project resources are openly available: the complete codebase, evaluation protocols, and analysis scripts at [https://github.com/juanjuanignacio/cardio_evals/], and the annotated cardiovascular QA database at [https://huggingface.co/datasets/Jialvareza/cardio_evaluations]. All analyses were run on a high-performance computing node equipped with 8 NVIDIA H100 NVL 94GB GPUs and CUDA 12.6. All API-based experiments were performed using the Ollama framework, a local runtime environment for serving large language models via API or direct execution. Reproducibility experiments were conducted using vLLM under batch and parallel local inference conditions.

## Supplementary Figures

**Supplementary Figure 1.**
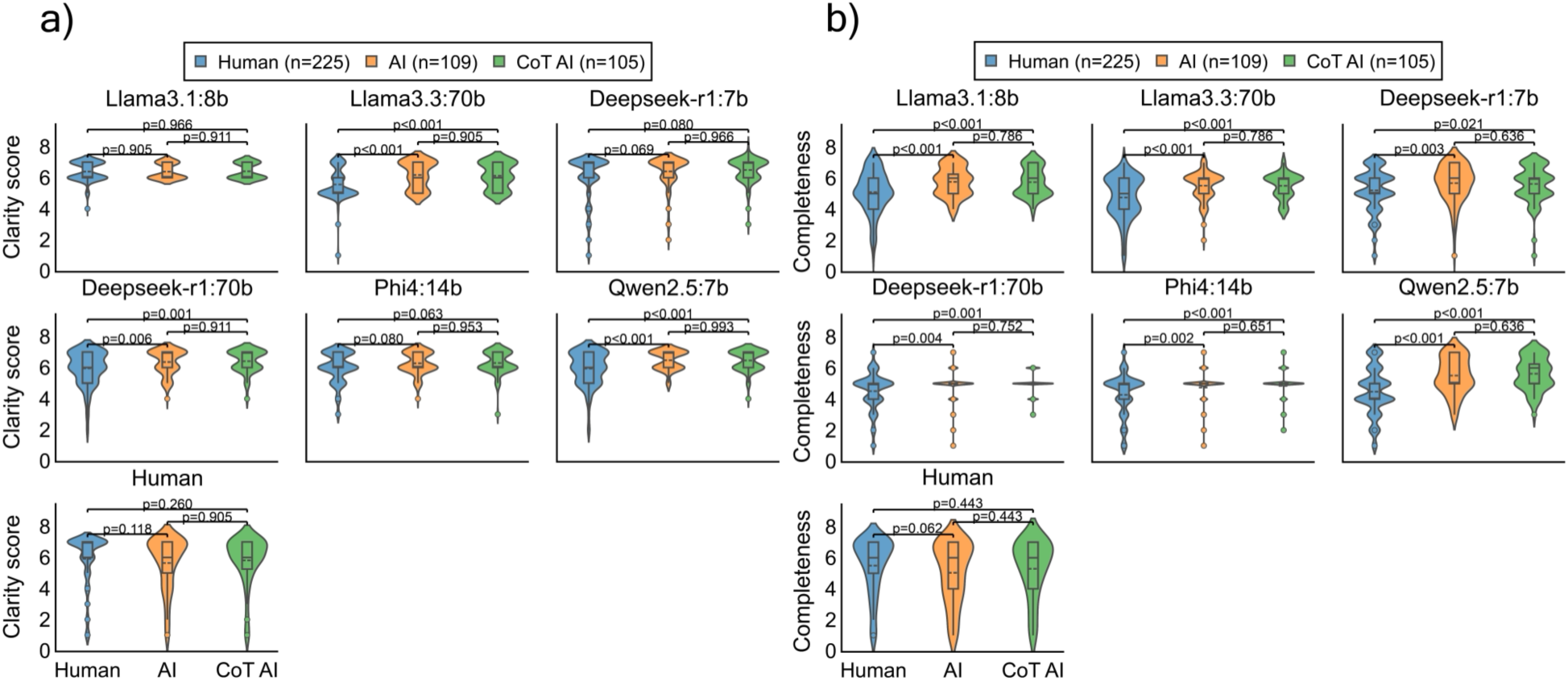
Comparison of Clarity (a) and Completeness (b) Scores Across Evaluator Models and Response Types. Violin plots showing the distribution of clarity scores assigned by different evaluator types to responses from three sources: human experts (blue), basic AI models (orange), and advanced AI models (green). Each panel represents a different evaluator model: Llama3.1:8b, Llama3.3:70b, DeepSeek-R1:7b, DeepSeek-R1:70b, Phi4, Qwen2.5, and human evaluators. Statistical comparisons employed Mann-Whitney U tests with Benjamini-Hochberg FDR correction for multiple testing.

**Supplementary Figure 2.**
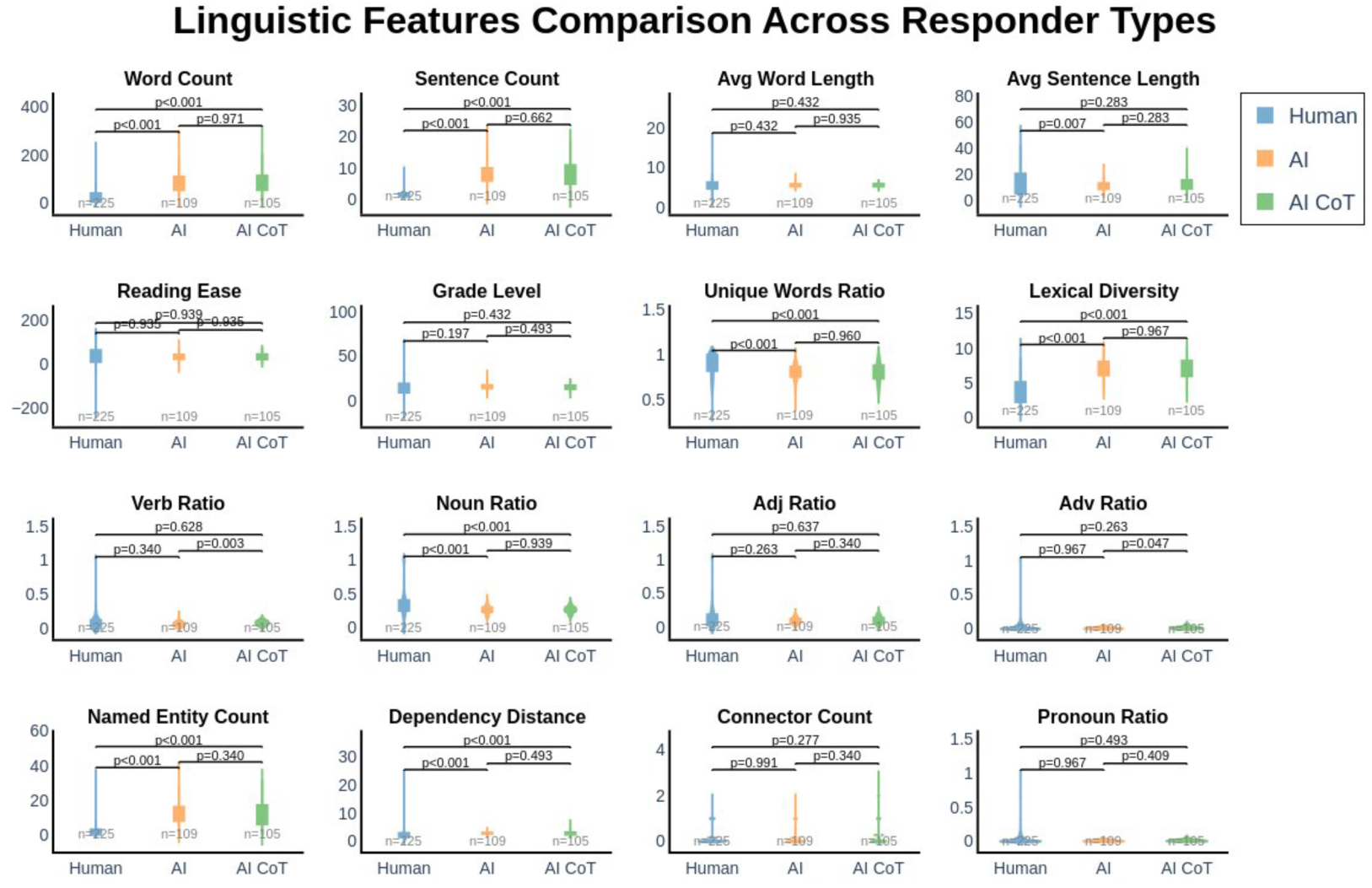
Distribution of linguistic features across response types after filtering ambiguous responses. Violin plots showing the distribution of 16 linguistic features for human expert (blue, n=125), standard AI (orange, n=109), and CoT AI (green, n=105) responses. Responses classified as “I don’t know” or equivalent were excluded from analysis. P-values correspond to pairwise Mann-Whitney U tests with Benjamini-Hochberg FDR correction applied across all 48 comparisons (16 features × 3 pairs). Human responses are significantly shorter than both AI types (word count and sentence count: Human vs AI p<0.001, Human vs CoT p<0.001), while standard AI and CoT AI do not differ in length (p=0.971 and p=0.662, respectively). Human responses show higher unique word ratio and lexical diversity compared to both AI types (p<0.001), but the two AI types do not differ from each other (p=0.960 and p=0.967). Standard AI responses are more noun-heavy than human responses (p<0.001) and contain more named entities (p<0.001), consistent with denser, more formal outputs. Dependency distance is significantly greater in both AI types compared to human responses (p<0.001), with no significant difference between standard and CoT AI (p=0.493). Notably, verb ratio differs significantly only between AI and CoT (p=0.003), with CoT showing higher verb usage. Reading ease distinguishes human responses from CoT AI (p<0.001) and shows borderline significance between standard AI and CoT (p=0.035), suggesting CoT responses are more complex. Most other features—including adjective ratio, connector count, and pronoun ratio—show no significant pairwise differences after multiple testing correction, though general trends suggest human responses employ a more conversational and personalized writing style.

**Supplementary Figure 3:**
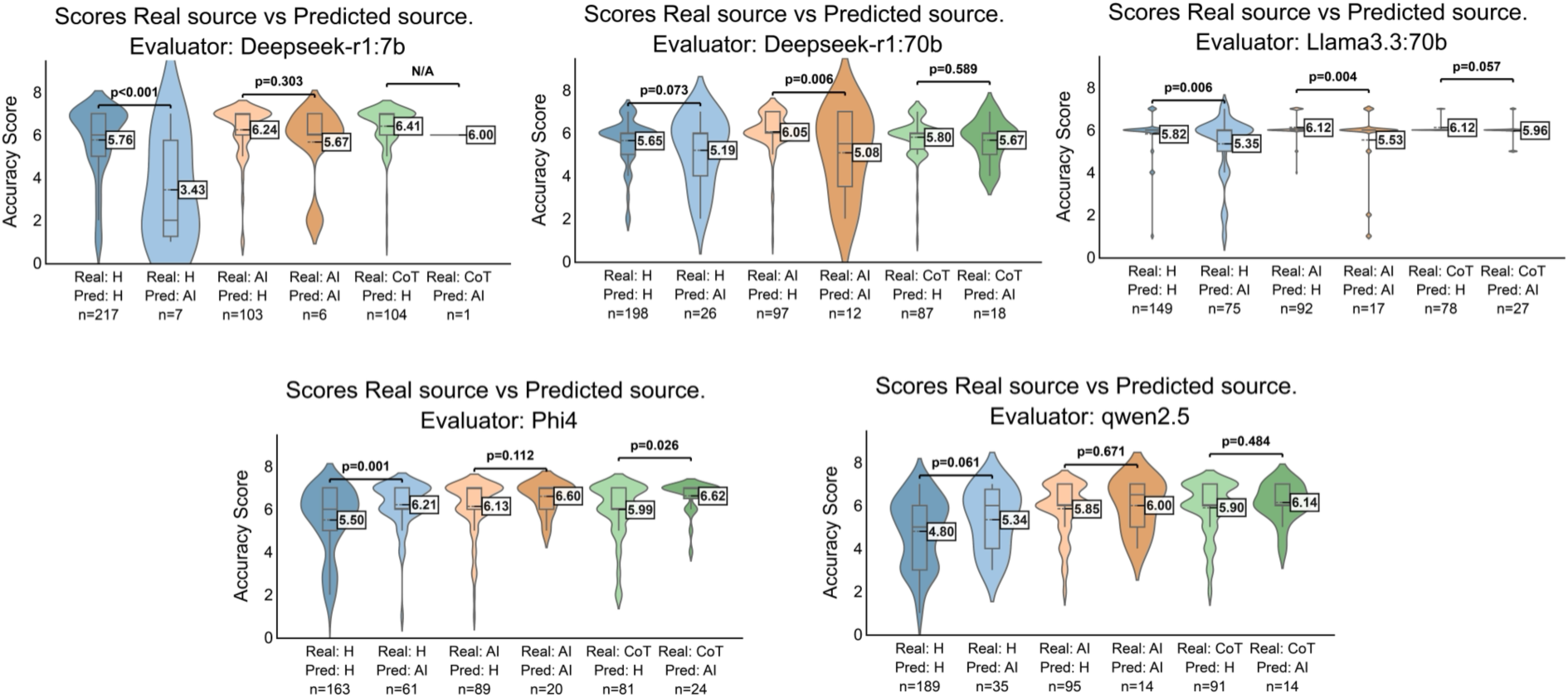
Distribution of accuracy scores given by different types of evaluators distinguishing between actual origin of responses and that predicted by the evaluator.

**Supplementary Figure 4.**
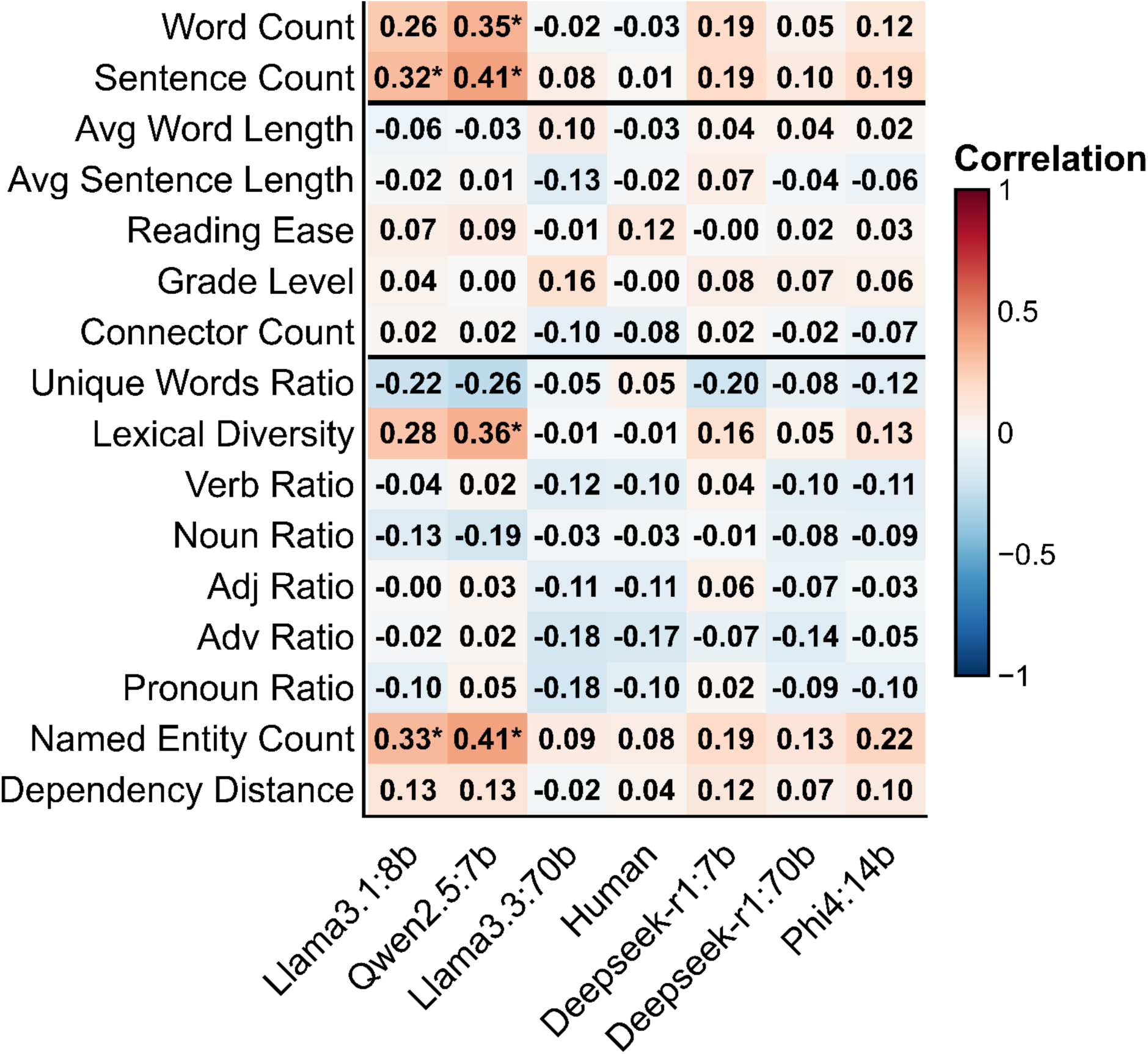
Spearman correlation between linguistic features and accuracy scores across evaluator types. Heatmap showing pairwise Spearman correlation coefficients between 16 linguistic features extracted from the responses and the accuracy scores assigned by each evaluator. Evaluators are clustered by similarity of rating patterns. Asterisks (*) denote statistically significant correlations based on 95% bootstrap confidence intervals not crossing zero.

## Data Availability

All data produced are available online at https://huggingface.co/datasets/Jialvareza/cardio_evaluations

## Notes

### Competing Interest Statement

The authors have declared no competing interest.

### Author Declarations

Ethics committee of Instituto de Salud Carlos III gave ethical approval for this work

### Summary of Updates

Author capital letters fixed; author order updated.

### Funding

- The CNIC is supported by the Instituto de Salud Carlos III (ISCIII), the Ministerio de Ciencia, Innovación y Universidades (MICIU) and the Pro CNIC Foundation, and is a Severo Ochoa Center of Excellence: CEX2020-001041-S
- This work was supported by JACARDI project, that has received funding from the EU4Health Program 2021-202.: 101126953

